# Bipolar anodal septal pacing with direct LBB capture preserves physiological ventricular activation better than unipolar left bundle branch pacing

**DOI:** 10.1101/2023.01.09.23284263

**Authors:** Karol Curila, Pavel Jurak, Frits Prinzen, Marek Jastrzebski, Petr Waldauf, Josef Halamek, Marketa Tothova, Lucie Znojilova, Radovan Smisek, Jakub Kach, Lukas Poviser, Hana Linkova, Filip Plesinger, Pawel Moskal, Ivo Viscor, Vlastimil Vondra, Pavel Leinveber, Pavel Osmancik

## Abstract

**Background:** Left bundle branch pacing (LBBP) produces delayed, unphysiological activation of the right ventricle. Using ultra-high-frequency electrocardiography (UHF-ECG), we explored how bipolar anodal septal pacing with direct LBB capture (aLBBP) affects the resultant ventricular depolarization pattern.

**Methods:** In patients with bradycardia, His bundle pacing (HBP), unipolar nonselective LBBP (nsLBBP), aLBBP, and right ventricular septal pacing (RVSP) were performed. Timing of local ventricular activation, in leads V1-V8, was displayed using UHF-ECG, and electrical dyssynchrony (e-DYS) was calculated as the difference between the first and last activation. Durations of local depolarizations were determined as the width of the UHF-QRS complex at 50% of its amplitude.

**Results:** aLBBP was feasible in 63 of 75 consecutive patients with successful nsLBBP. aLBBP significantly improved interventricular dyssynchrony (−9 ms (−12;−6) vs. −24 ms (−27;−21),), p < 0.001) and shortened local depolarization durations in V1–V4 (mean differences −7 ms to −5 ms (−11;−1), p < 0.05) compared to nsLBBP. Both aLBBP and HBP caused similar absolute levels of interventricular dyssynchrony (e-DYS −9 ms (−12; −6) vs. 10 ms (7;14); however, local depolarization durations in V1–V2 during aLBBP were longer than HBP (differences 5–9 ms (1;14), p < 0.05, with local depolarization duration in V1 during aLBBP being the same as during RVSP (difference 2 ms (−2;6), p = 0.52).

**Conclusion:** Although aLBBP significantly improved interventricular synchrony and depolarization duration of the septum and RV compared to unipolar nsLBBP, the resultant ventricular depolarization was still less physiological than during HBP.

## Introduction

Left bundle branch pacing (LBBP) is an emerging treatment strategy for patients with bradycardia (1). As shown recently, using an Ultra-high-frequency ECG (UHF-ECG), LBBP is less physiological than His bundle pacing (2); however, it has better sensing values, pacing thresholds, and offers favorable clinical outcomes making it a more attractive option in many cases.

During LBBP, left ventricular (LV) depolarization precedes right ventricular (RV) activation, creating interventricular dyssynchrony. The delayed RV activation results in a pseudo-right bundle branch morphology in lead V1, which is one of the proposed markers of a successful left bundle branch capture (3). The lead tip is placed in the left septal subendocardial area during the implant procedure, which often situates the ring electrode in contact with the right septal endocardium. As shown recently, this allows both the cathode and anode of pacing leads to simultaneously capture the right and left septal subendocardial areas during bipolar pacing at higher pacing outputs (4, 5). Such pacing can lead to more balanced ventricular activation and eliminate the pseudo-right bundle branch morphology in V1. However, the exact impact of this type of pacing on ventricular depolarization has never been fully described.

This project aimed to use UHF-ECGs to study how bipolar anodal septal capture during LBBP changes ventricular depolarization patterns compared to unipolar capture of the LBB, His bundle, and right ventricular septal pacing (RVSP) in patients with bradycardia.

## Methods

In this prospective study, all consecutive patients with an indication for pacemaker implantation due to bradycardia and successful LBB capture were included. The study was approved by the Ethics Committee of the Faculty Hospital Kralovske Vinohrady, and all patients signed informed consent before enrollment.

Implant procedures started with mapping the His bundle using a SelectSecure™ lead (model 3830, 69 cm, Medtronic Inc., Minneapolis, MN), delivered through a fixed-curve sheath (C315 HIS, Medtronic, Minneapolis, MN). The His bundle signal was identified, and its nonselective capture (HBP) was confirmed using decremental output pacing (6). The lead was then moved towards the right ventricle and screwed deep into the septum to obtain a position on the left side of the interventricular septum. The targeted area for lead screwing was 1.5–2 cm below the level of the His bundle region or vertex of the tricuspid valve visualized using contrast injection through the C315 HIS (3, 7) and preferentially with a normal paced heart axis. Lead screwing was occasionally interrupted, and unipolar pacing with a 5 V output at 0.5 ms was performed. Once the paced QRS morphology demonstrated a terminal r/R morphology in lead V1 or significant narrowing compared to RVSP, decremental output pacing was then performed to identify nonselective left bundle branch capture (nsLBBP). nsLBBP capture was proved when a transition to selective left bundle branch capture (sLBBP) or myocardial LV septal capture during decremental output pacing was seen (1, 3). After that, the alligator clip was moved to pace from the ring electrode of the pacing lead, and unipolar pacing with outputs up to 5 V at 0.5 ms was performed. When right septum capture was successful (i.e., RVSP was feasible), both alligator clips were connected to the pacing lead (tip as cathode, ring as anode), and bipolar pacing was performed with a maximum pacing output of 10 V at 0.5 ms. The pacing output was then slowly reduced, and changes in the intracardiac electrogram (EGM) signal and QRS morphology were carefully inspected. Once the EGM signal became more ‘distinct,’ and the QRS morphology in V1 showed progression in the amplitude of the late r/R, the transition from aLBBP to nsLBBP was identified, and the threshold value was noted (Figure 1). UHF-ECG recordings were performed using the unipolar lead connection during HBP, nsLBBP, RVSP, and the bipolar lead connection during aLBBP. The pacing outputs during HBP and RVSP were 5 V at 0.5 ms; pacing outputs during nsLBBP and aLBBP were the same (usually 5 V at 0.5 ms).

**Figure 1:**
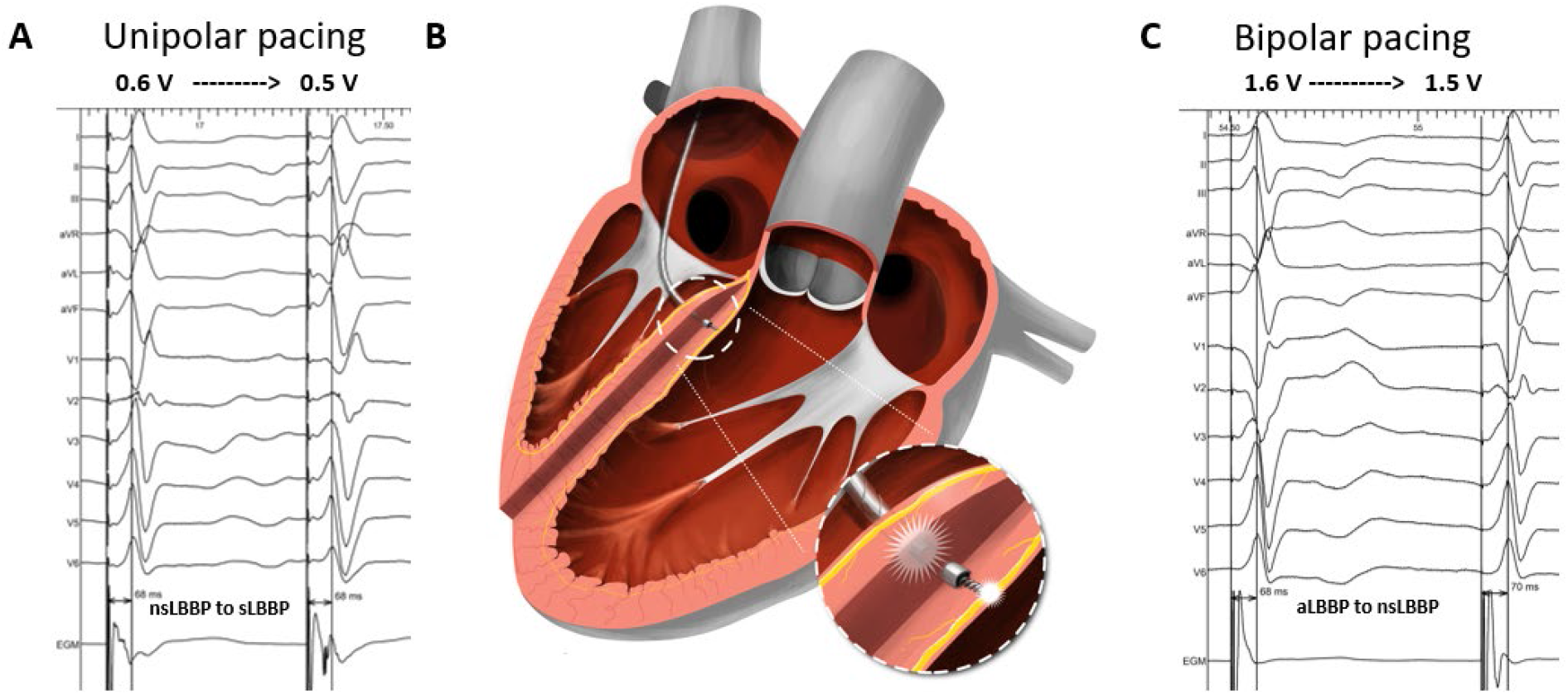
Identification of nsLBBP during unipolar pacing in panel A (nsLBBP changed to sLBBP during decremental output pacing) and anodal septal LBB capture (aLBBP) transitioning to nsLBBP at 1.6 V at 0.5 ms during bipolar pacing in panel C. Visualization of bilateral septal capture with tip and ring electrode of the pacing lead during bipolar aLBBP is shown in panel B.

### UHF-ECG data acquisition and analysis of other measured parameters

A VDI monitor (Ventricular Dyssynchrony Imaging monitor, ISI Brno, Cardion, FNUSA, CZ, 2018) was used to record and analyze the 5 kHz 14-lead ECG signals with a three nV resolution and a frequency range of 1.5 kHz. Standard V1-V8 chest lead positions were used, except for lead V1, which was moved from the fourth to the fifth right parasternal intercostal space to obtain better signals from the lateral RV wall. UHF-ECG data for all captures were collected during 2–3 minutes of VVI pacing at 110 beats/min. Signal processing and UHF-ECG map construction are described elsewhere (8). Median amplitude envelopes were computed in 16 frequency bands (150–1000 Hz) for each chest lead. The broad-band QRS complex (UHF-QRS) was constructed as the average of the 16 normalized median amplitude envelopes and displayed as a colored map for V1–V8 leads (ventricular depolarization map). Local activations were calculated as the center of mass of the UHF-QRS above the 50 percent threshold of the baseline-to-peak amplitude for each chest lead (V1-V8). Local depolarization durations under leads V1–V8 were computed as the UHF-QRS duration at 50 percent of its amplitude. Interventricular electrical dyssynchrony, i.e., e-DYS, was calculated as the difference between the first and last activation on a UHF-ECG map (Figure 2). A positive e-DYS indicates delayed LV activation, and a negative e-DYS indicates delayed RV activation.

**Figure 2:**
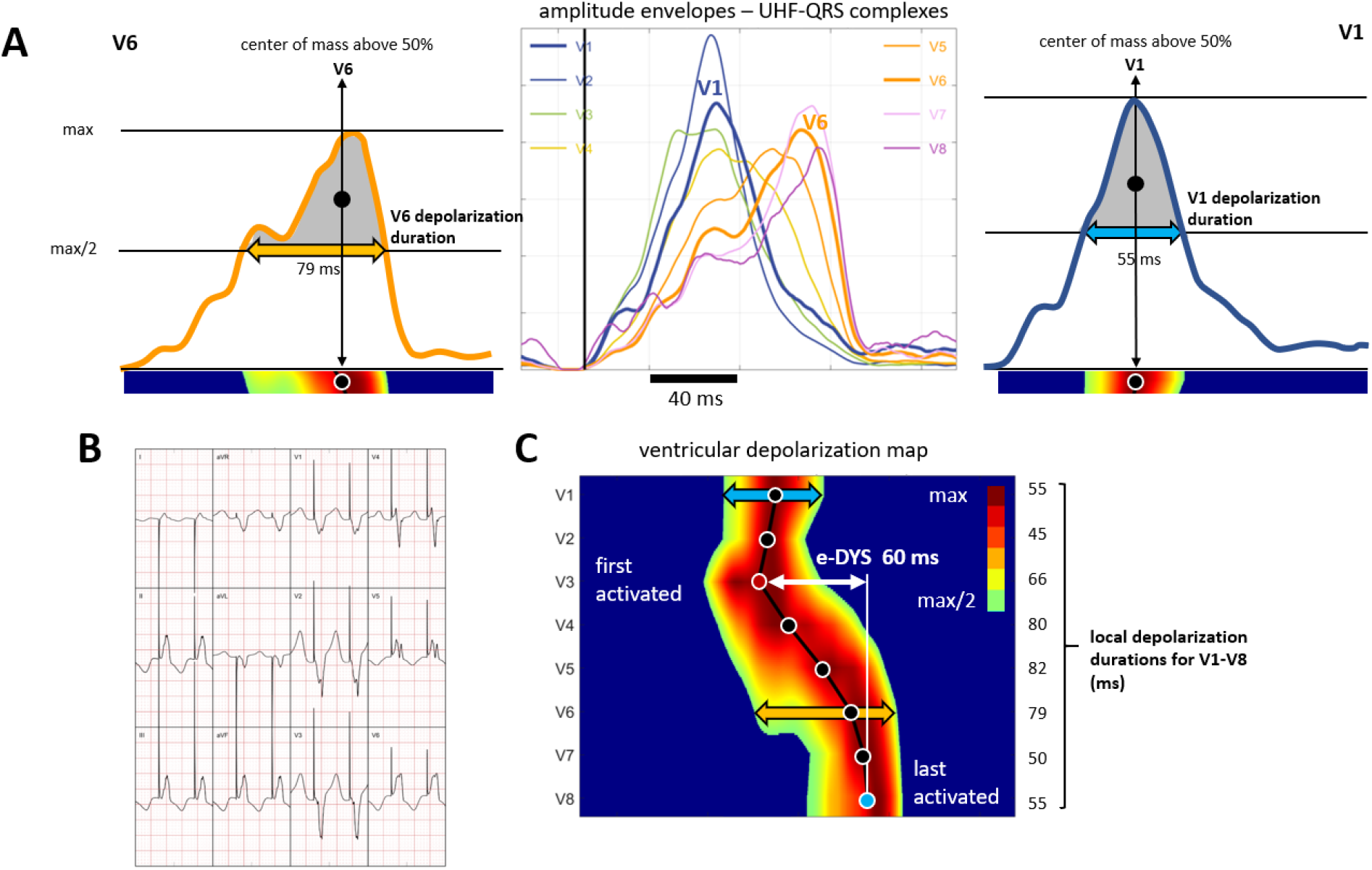
Presentation of amplitude envelopes of UHF-QRS complexes, local activations, and local depolarization durations calculation; patient with RVSP. (Panel A) amplitude envelopes of V1-V8 UHF-QRS complexes (in the middle), local activations (solid circles) computed as the center of mass above the 50 percent threshold of the baseline to peak amplitude, and local depolarization durations determined as the depolarization duration at the 50 percent threshold of the baseline to peak amplitude for leads V6 (left) and V1 (right). (Panel B) 12-lead ECG. (Panel C) ventricular depolarization map with visualization of the timing of local activations (solid circles) in leads V1-V8, electrical interventricular dyssynchrony e-DYS, and local depolarization duration values. For details, see Jurak, P. et al. (2020).

Global QRS durations (QRSd) were measured using an electrophysiology system (Labsystem Pro, Boston Scientific, USA) from the earliest to the last deflection in any of the 12 leads (100 mm/s sweep speed, 16 x digital augmentation, average measurement from two consecutive beats). During pacing, the beginning of the QRS was measured from (1) the pacing artifact (QRSd) and (2) the first deflection identified after the pacing artifact (actual QRSd). The paced V5 RWPT was measured from the pacing artifact to the maximum positive QRS amplitude in lead V5. Echocardiography was performed in the left supine position using standard projections. LVEF was estimated from parasternal and apical projections; the diameter of the RV and LV and septal thickness were measured, during diastole, using the parasternal long axis projection in 2D mode.

## Statistics

An exploratory data analysis was performed for all parameters. Unpaired comparisons of continuous and categorical variables were made using the unpaired t-test and Chi-square test. Paired and repeated measurement comparisons were made using a linear mixed effect model (LMEM) and the Tukey multiple comparison test. The results of these models are presented as means with 95% confidence intervals and comparisons as mean differences with 95% confidence intervals and p-values (Figures 3,4,5,6, Supplementary Figure 1). A p-value < 0.05 was considered statistically significant. RStudio version 1.2.1335 with R version 3.6.1 was used to perform the statistical analyses. The LMEM was calculated using lme4 version 1.1–21. If not specified, all values are shown as means (95% CI).

**Figure 3:**
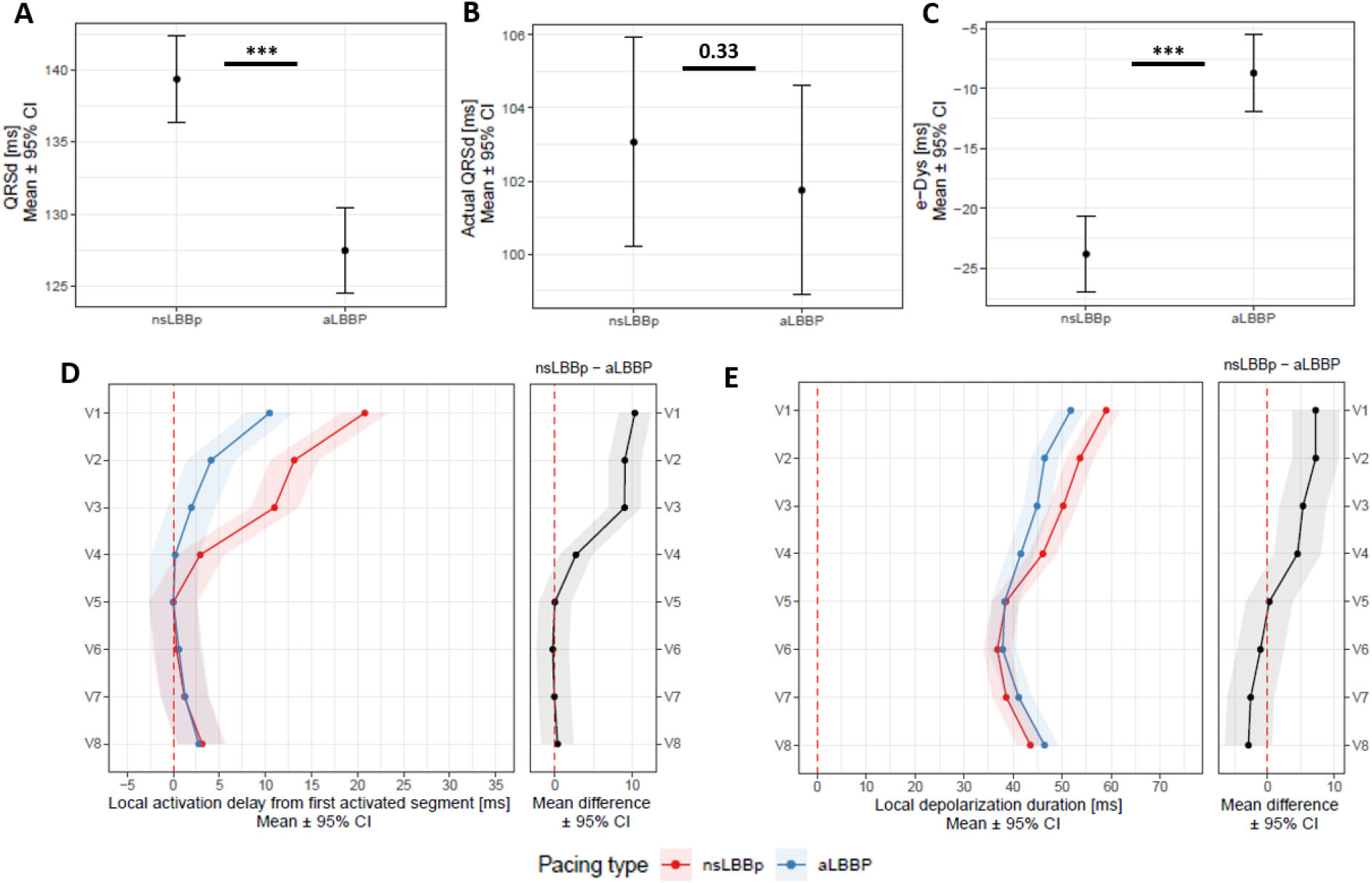
Panel A: QRSd, actual QRSd (Panel B), e-DYS (Panel C), local activations in V1-V8 (first activated segment was placed at 0 ms) (Panel D), and in Panel E, local depolarization durations in V1–V8 between nsLBBP and aLBBP. *** p < 0.001

**Figure 4:**
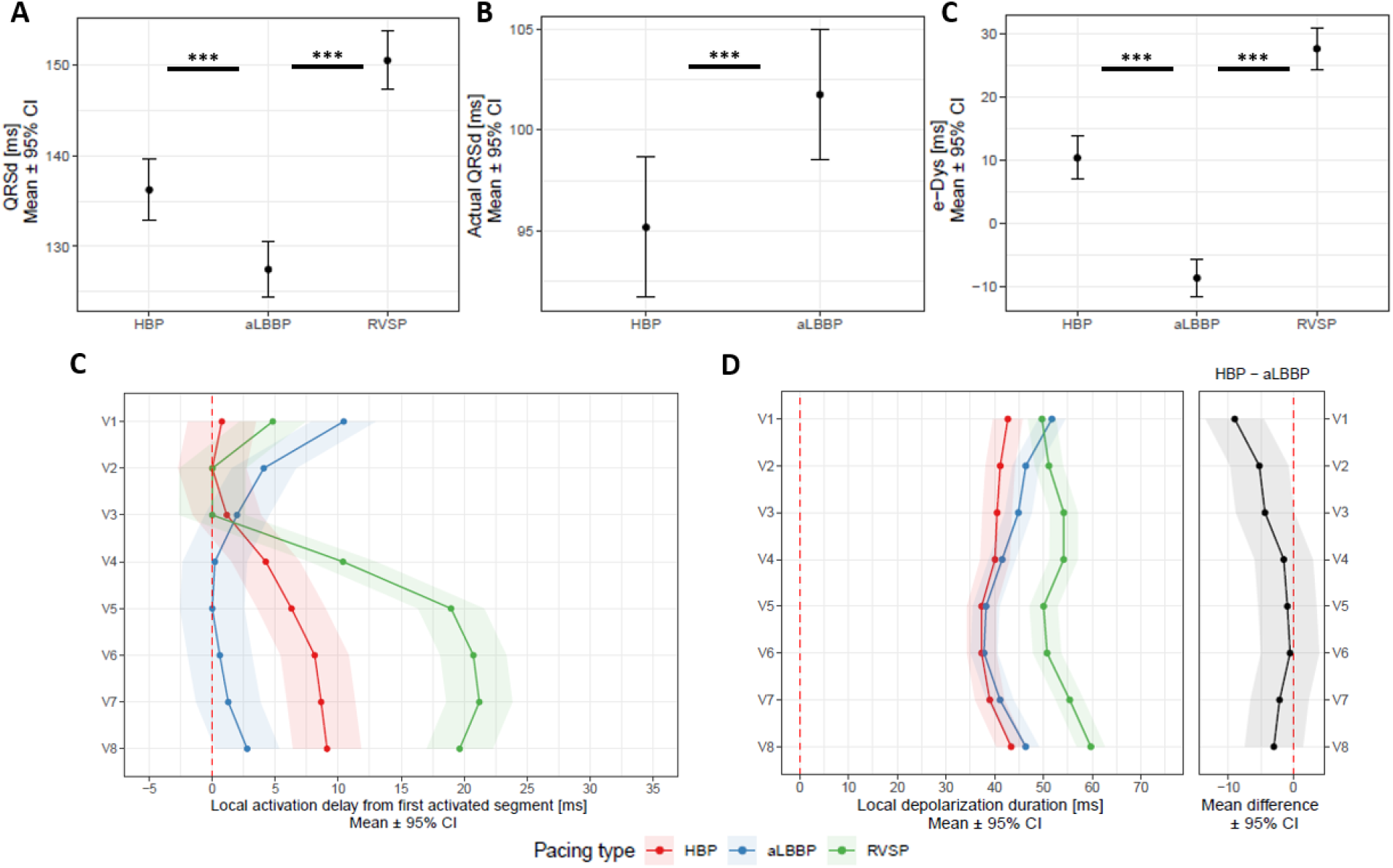
Panel A: QRSd, actual QRSd (Panel B), e-DYS (Panel C), local activations in V1-V8 (first activated segment was placed at 0 ms) (Panel D), and in Panel E, local depolarization durations for V1–V8 between HBP, aLBBP, and RVSP. *** p < 0.001

**Figure 5:**
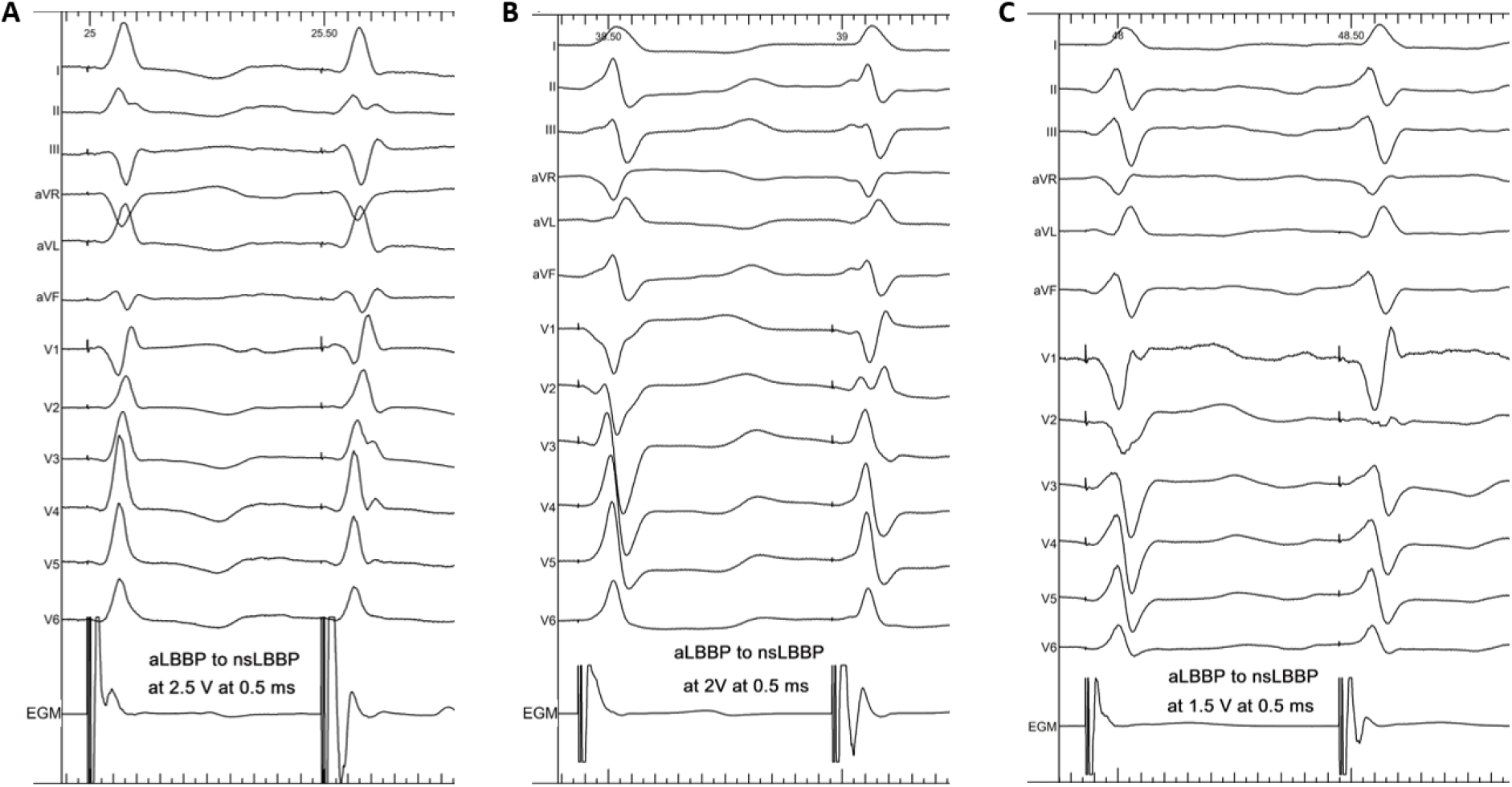
Change from aLBBP with a late r/R morphology to nsLBBP (Panel A), aLBBP with a QS morphology to nsLBBP (Panel B), and aLBBP with Qrs morphology to nsLBBP (Panel C).

**Figure 6:**
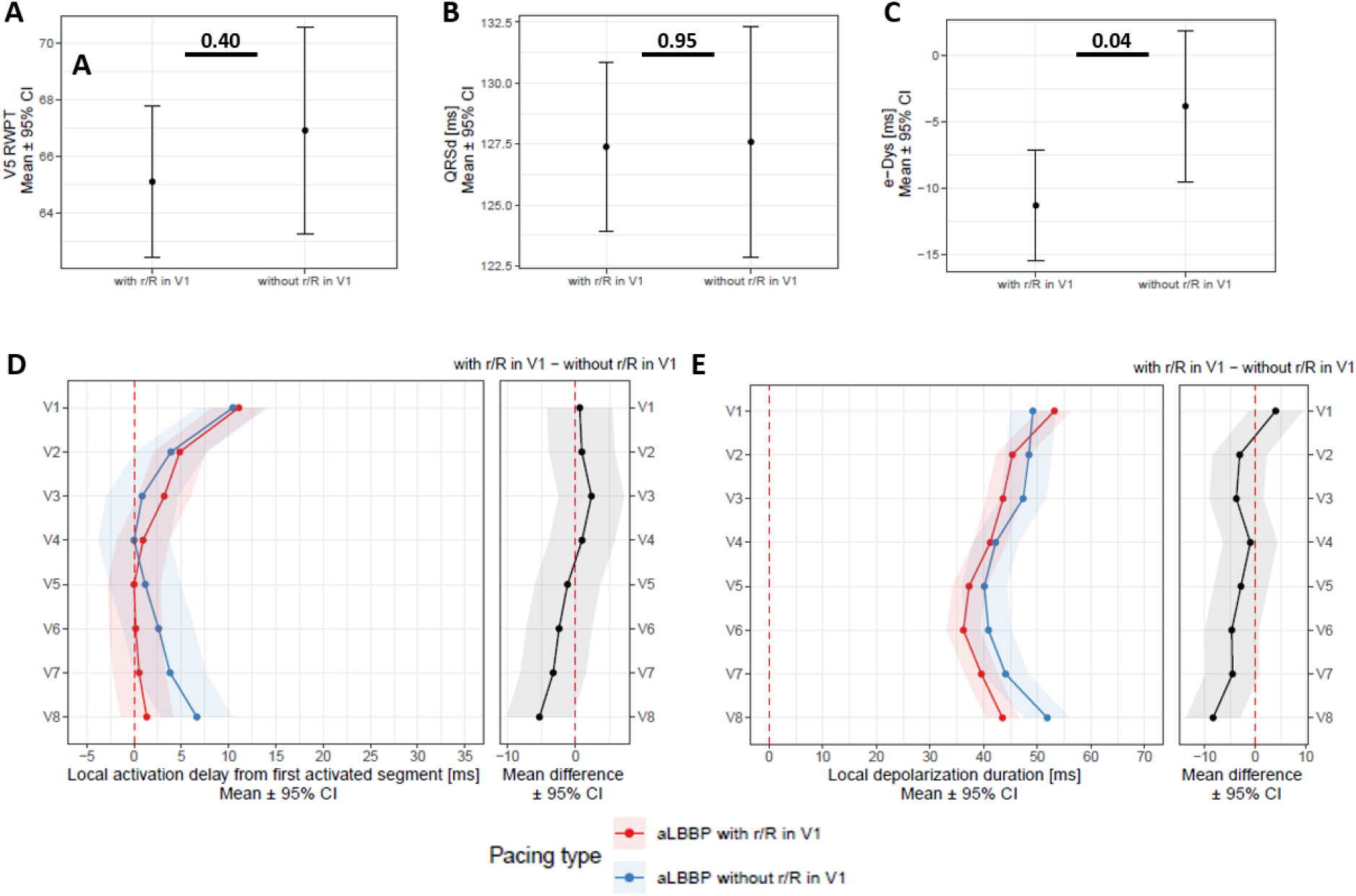
Panel A: V5RWPT, QRSd (Panel B), e-DYS (Panel C), local activations in V1-V8 (first activated segment was placed at 0 ms) (Panel D), and in Panel E, local depolarization durations in V1–V8 between aLBBP with r/R and without r/R in lead V1.

## Results

Of 116 consecutive patients with bradycardia, aLBBp was successful in 63 of 75 patients with unipolar nsLBBP demonstrated during an implant procedure using the decremental pacing output. Characteristics of patients with aLBBP are shown in Table 1. The average pacing threshold leading to aLBBP during bipolar pacing was 2.7 ± 0.9 V at 0.5 ms. In all 63 patients, both nsLBBP and aLBBP were recorded, all together with 51 nonselective His bundle captures (HBP) and 56 RVSP.

**Table 1:**
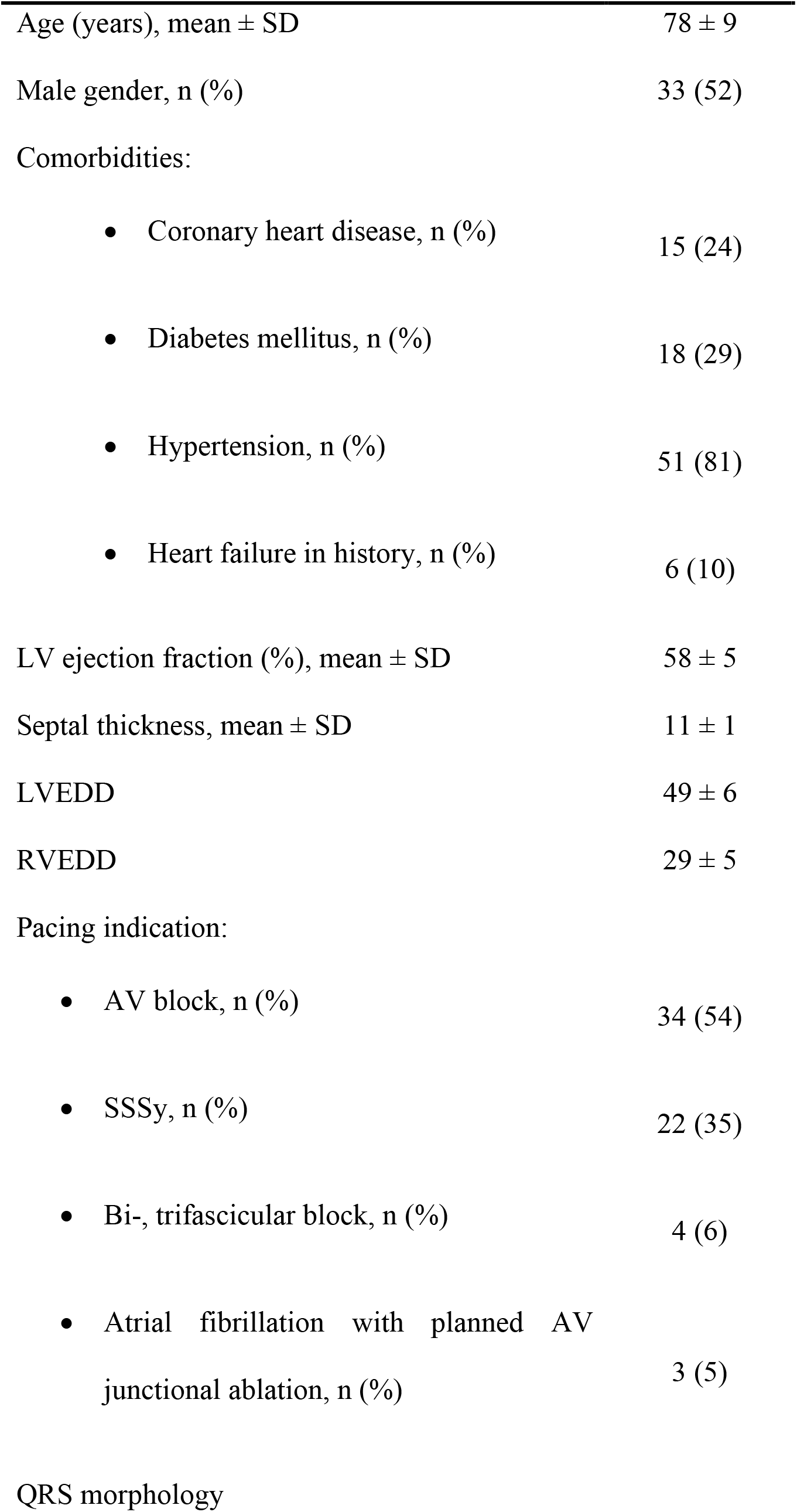
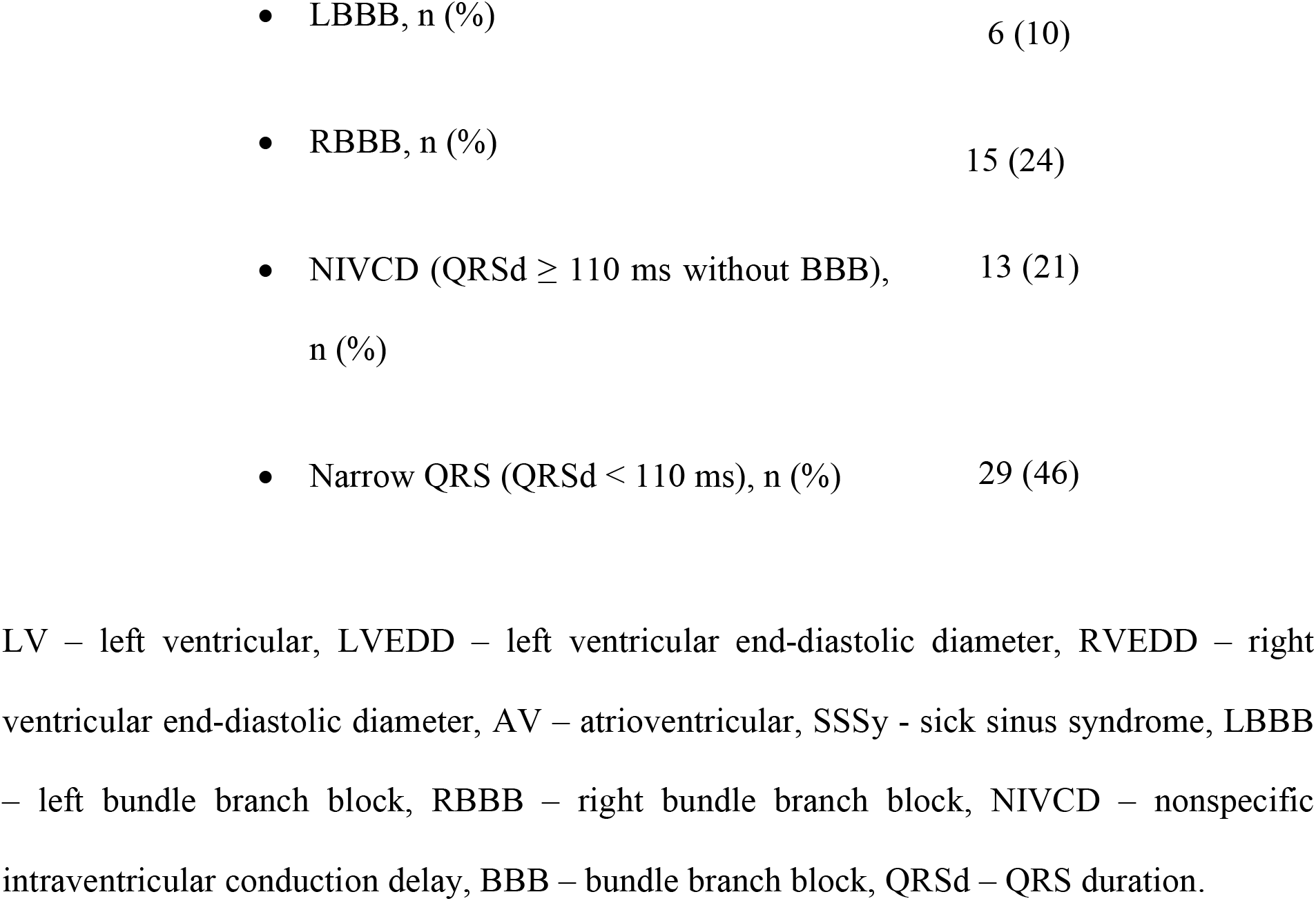
Clinical characteristics of patients with aLBBP during implant procedures.

aLBBP had shorter R-wave peak time in V5 (V5RWPT) than nsLBBP (66 ms (64;68) vs. 67 ms (65;69), p = 0.01). The QRSd measured from the pacing stimulus during nsLBBP was longer than during aLBBP, but the actual QRSd values were the same, which was a consequence of earlier RV activation during aLBBP, i.e., the isoline after the pacing artifact during aLBBP was shortened by −11 ms (12;-9), p < 0.001 compared to nsLBBP. The UHF-ECG ventricular activation sequence during both captures was the same under leads V5–V8. However, during aLBBP activations under V1–V4 were less delayed compared to nsLBBP, which led to e-DYS shortening. Similarly, no difference in local depolarization durations under the leads associated with LV lateral wall was observed, but their values in V1–V4 were significantly shorter during aLBBP than during nsLBBP (Figure 3).

To understand how aLBBP differs from physiological ventricular activation (HBP) and unphysiological ventricular activation during RVSP, we compared them to each other. Both HBP and aLBBp preserved the interventricular dyssynchrony at the same absolute level; however, HBP led to a right to left depolarization pattern while aLBBP led to a left to right depolarization pattern (e-DYS 10 ms (7;14) for HBP vs. −9 ms (−12; −6) for aLBBP, p < 0.001). QRSd was the shortest during aLBBP, but HBP had shorter actual QRSd and produced the most physiological activation of both ventricles with the shortest local depolarization durations in V1–V8. Although aLBBP had the same local depolarization durations in V4–V8 as HBP, V1–V3 values were significantly longer during aLBBP, and depolarization duration in V1 during aLBBP was the same as during RVSP. RVSP led to the longest e-DYS and local depolarization durations in V2–V8 of the studied ventricular captures (Figure 4).

Fifty-nine of 63 nsLBBP had a pseudo-right bundle branch block morphology with late r/R in V1 during pacing. During aLBBP, late r/R in lead V1 was seen in 41 patients. In the others without the r/R morphology in V1 during aLBBP, either a QS (n = 17) or a Qrs (n = 5) V1 morphology was observed (Figure 5). Comparing aLBBP with and without the r/R morphology in V1, we found no difference between V5RWPT and QRSd (Figure 6, Panels A, B). However, those without r/R in lead V1 had later local activation and longer depolarization durations under lead V8 (Figure 6, Panels D, E) and smaller interventricular dyssynchrony compared to aLBBP with late r/R (Figure 6, Panel C). In addition, a positive e-DYS was seen in 12 of 22 (55%) aLBBP without r/R compared to 5 of 41 (12%) for those with the r/R morphology in V1, p = 0.001.

To understand which patients with nsLBBP will not have a late r/R in V1 during aLBBP, we compared nsLBBP that transited to aLBBP without the r/R morphology to those that transited to aLBBP with r/R in lead V1. We did not find any significant difference in age, gender distribution, presence of diabetes mellitus, coronary artery disease, or hypertension. Also, LVEF, septal thickness, LVED, and RVED diameters, and the presence of BBB, IVCD, or narrow QRS complexes with QRSd below 110 ms during spontaneous rhythm were the same in both groups (Supplementary Table 1). The only differences were the timing of V8 local activation from the pacing artifact, which during nsLBBP transitioning in aLBBP with the r/R morphology was earlier by −7 ms (−13;0), p = 0.04 and in local depolarization durations in V6–V7, which were shorter for −7 (−14;0), p = 0.04 and p = 0.03 respectively compared to nsLBBP terminating in aLBBP without the r/R morphology in V1 (Supplementary Figure 1).

## Discussion

Our study showed that anodal septal pacing with LBB capture was feasible in most patients with bradycardia and successful unipolar LBB pacing, with an average capture threshold of 2.7 V at 0.5 ms. aLBBP improved interventricular synchrony without worsening LV lateral wall depolarization compared to unipolar LBB capture. aLBBP preserved interventricular synchrony and physiological LV activation to the same degree as HBP but led to prolonged depolarization durations under the leads placed above the septum and right ventricle, with the depolarization duration in V1 being the same as during right ventricular septal capture. aLBBP without the r/R morphology led to less interventricular dyssynchrony than aLBBP with late r/R morphology in the lead V1. The reason for the improved interventricular synchrony was delayed and prolonged activation of myocardial segments under V8, which was a consequence of longer electrical wave-front propagation within the LV lateral wall already present during unipolar LBB capture.

### aLBBP vs. nsLBBP

Although nsLBBP leads to similar physiological LV activation as HBP, it creates left-to-right interventricular dyssynchrony (2). However, specific lead design and deep septal placement during LBBP made it possible to simultaneously pace the ring and tip electrode to improve ventricular synchrony. The effect of anodal bipolar LBBP on QRSd, interventricular synchrony, and capture thresholds was recently described in two studies (4, 5). Both reported the average pacing threshold leading to aLBBP being below 3V at 0.4 ms (2.7 V in the Lin and 2.5 in the Wu study) and a success rate of 61% (22 of 36 patients) and 87% (65 of 75 patients), respectively. They also observed that this type of pacing, compared to unipolar LBBP, led to QRSd narrowing and an improvement in interventricular dyssynchrony, based on time delay measurements between V6RWPT and V1RWPT. The higher success rate of aLBBP in our study compared to Lin’s study is very likely due to the different septal thicknesses in the studied populations. The septa of our patients were thicker, which allowed the ring electrode to be in contact with or within the RV septal endocardium in most patients. We observed aLBBP capture thresholds at levels (2.7 ± 0.9 V at 0.5 ms) similar to the studies mentioned above and improvements in QRSd and interventricular dyssynchrony during aLBBP. We also demonstrated that performing aLBBP shortens the depolarization of the ventricular segments under V1-V4 compared to unipolar LBBP. Moreover, it does not worsen LV lateral wall activation compared to HBP and unipolar nsLBBP, the first of which has already been shown to be the most physiological type of ventricular pacing (2, 9).

### aLBBP vs. HBP and RVSP

We have already shown that the pacing of the RV septum in its inflow tract caused the least LV lateral wall delay of all RV pacing locations except for ventricular HB or proximal RBB pacing (9). The present work shows that both HBP and aLBBP reduce the absolute value of interventricular dyssynchrony by approximately three times compared to RVSP. Moreover, local depolarization durations in the ECG leads associated with the LV were approximately 25% shorter during aLBBP and HBP than RVSP. This is a consequence of fast LV activation due to the direct excitation of LBB conductive tissue. Although aLBBP was associated with more physiological ventricular depolarization than RVSP and unipolar LBBP, its local depolarization durations under V1-V3 remained longer than during HBP. Their prolongation results from slower septal and RV activation during aLBBP compared to HBP, during which the septum and RV are activated more rapidly due to direct capture of the right bundle branch. The impetus for developing new pacing strategies such as HBP and LBBP was that dyssynchronous ventricular activation during RV pacing worsens clinical outcomes in some patients. Although LBBP showed it could overcome most HBP shortcomings (10), the clinical significance of resultant interventricular dyssynchrony during LBBP is unclear. Currently, no data indicate that the effects of LBBP-induced delays in RV activation on clinical outcomes in bradycardia patients are detrimental. Data from one observational trial showed that both types of left septal pacing (LBBP and LVSP) were clinically superior to RV myocardial pacing (11), although the rates of unipolar vs. bipolar anodal captures were not reported.

### The influence of aLBBP on RV and LV activation and their depolarization interplay

The formation of late r/R in V1 during nsLBBP reflects that after pacing, the LV activates rapidly through the His-Purkinje system, but RV activation is delayed due to trans-septal wave-front propagation and slower conduction within the RV. During aLBBP, RV activation is less delayed because both LV and RV septal areas are captured simultaneously. Moreover, as our data show, RV and septal activation is not only less delayed during aLBBP, but their activation is also more physiological than during unipolar LBBP, with shorter depolarization durations under V1-V4. However, compared to HBP, activations under V1 and V2 were prolonged during aLBBP, and under V1, they were practically the same during aLBBP and RVSP. This can be explained by differences in electrical wave-front propagation within the RV during the studied captures. During aLBBP and RVSP, the RV septum is captured immediately. The electrical wave-front spreads through subendocardial connective tissue faster [12] than the slower cell-to-cell propagation during nsLBBP. However, HBP still offers more physiological RV activation since it activates the RV through RBB fibers.

As our results show, the disappearance of the r/R in V1 during aLBBP is a marker of better ventricular synchrony and longer depolarization durations in ventricular segments under V8 compared to captures where the r/R in V1 is present. Our data suggest that this type of balanced ventricular activation during aLBBP is very likely the result of the underlying latency of LV lateral wall activation, which is already present during unipolar nsLBBP, and not a specific effect of aLBBP with simultaneous capture of RBB and LBB fibers. Although capturing both branches during aLBBP is a tempting goal, it would be difficult in most patients because the paths the fibers follow along the left, and right septal surfaces are different for most of the course (12).

## Conclusion

Our study showed that in most patients with bradycardia, delayed RV activation and longer septal and RV depolarization during nonselective LBB pacing could be improved using simultaneous pacing of both septa during ‘anodal’ bipolar pacing with LBB capture. This type of pacing significantly improved interventricular dyssynchrony, septal, and RV depolarization durations compared to unipolar LBB pacing; moreover, it uses pacing outputs that are feasible in most patients with pacemakers implanted for bradycardia. Although aLBBP produced more physiological ventricular depolarization than unipolar LBBP, it was still less physiological than HBP. HBP led to faster depolarization of the RV, with RV depolarization during aLBBP being just as fast as during right ventricular septal pacing. The disappearance of the r/R pattern in V1 during aLBBP is a marker of more balanced ventricular activation, which is a consequence of longer electrical wave-front propagation within the LV during unipolar LBB capture rather than simultaneous capture of both Tawara branches.

### Limitations

Although we demonstrated that aLBBP provides more physiological ventricular electrical activation, we did not show that it results in a better hemodynamic response or less mechanical dyssynchrony than nsLBBP. Also, the small differences in the measured values of electrical dyssynchrony have unknown clinical relevance, and their significance, if any, needs to be confirmed in prospective clinical trials. This study was performed during actual implant procedures. UHF-ECG measurements were taken immediately after the lead was placed in predefined positions and after the type of ventricular capture was confirmed. We cannot rule out that procedure-related damage to conductive and myocardial tissue could have influenced the paced ventricular depolarization patterns. In three patients, poor QRS signal quality prevented the construction of UHF-ECG maps; therefore, these patients were excluded from the study.

## Data Availability

All data produced in the present study are available upon reasonable request to the authors.

## Acknowledgments

This paper was supported by the Charles University Research Program Cooperatio – Cardiovascular Science (KC), Ministry of Health of the Czech Republic, grant number NU21-02-00584 (KC), National Institute for Metabolic and Cardiovascular Research"CarDia” (project nr. LX22NPO5104) (KC), by the CAS project RVO:68081731 (PJ) and the European Regional Development Fund - Project ENOCH No.CZ.02.1.01/0.0/0.0/16_019/0000868 (PL).

## Data availability statement

The data that support the findings of this study are available on request from the corresponding author. The data are not publicly available due to privacy or ethical restrictions.

---

**Supplementary Table 1:** A comparison of patients with nsLBBP transitioning to aLBBP with the r/R morphology to those transitioning to aLBBP without the r/R morphology in V1.

|  | nsLBBP<br>transitioning to<br>aLBBP with late<br>r/R morphology in<br>V1<br>n = 41 | nsLBBP<br>transitioning to<br>aLBBP without the<br>r/R morphology in<br>V1<br>n = 22 | p value |
| --- | --- | --- | --- |
| Age (years), mean ± SD | 78 ± 10 | 79 ± 6 | 0.54 |
| Male gender, n (%) | 23 (56) | 10 (45) | 0.42 |
| Comorbidities: |  |  |  |
| • Heart failure, n (%) | 5 (12) | 1 (5) | 0.32 |
| • Coronary heart disease, n (%) | 8 (20) | 7 (32) | 0.36 |
| • Diabetes mellitus, n (%) | 10 (24) | 8 (36) | 0.31 |
| • Hypertension, n (%) | 31 (76) | 20 (91) | 0.12 |
| LV ejection fraction (%), mean ± SD | 59 ± 5 | 58 ± 4 | 0.52 |
| Septal thickness, mm, mean ± SD | 10.8 ± 1 | 11.0 ± 1 | 0.41 |
| LVEDD, mm, mean ± SD | 49 ± 6 | 49 ± 5 | 0.80 |
| RVEDD, mm, mean ± SD | 29 ± 4 | 29 ± 5 | 0.82 |
| Pacing indications: |  |  |  |
| • AV block, n (%) | 17 (41) | 7 (32) | 0.45 |
| • SSSy, n (%) | 9 (22) | 9 (41) | 0.11 |
| • Bi-, trifascicular block, n (%) | 2 (5) | 1 (5) | 0.95 |
| • Atrial fibrillation with planned<br>AV junctional ablation, n (%) | 2 (5) | 0 (0) | NA |
| QRS morphology |  |  |  |
| • LBBB, n (%) | 4 (10) | 2 (9) | 0.93 |
| • RBBB, n (%) | 9 (22) | 5 (23) | 0.94 |
| • IVCD, n (%) | 5 (12) | 3 (14) | 0.87 |
| • Narrow QRS, n (%) | 23 (56) | 12 (54) | 0.91 |
| e-DYS, ms, mean ± SD | -23 | -25 | 0.73 |
| QRSd, ms, mean ± SD | 139 | 141 | 0.56 |
| V5RWPT, ms, mean ± SD | 66 | 69 | 0.34 |

LVEDD – left ventricular end-diastolic diameter, LVESD – left ventricular end-systolic diameter, all other abbreviations were previously used in the text.

**Supplementary Figure 1:**
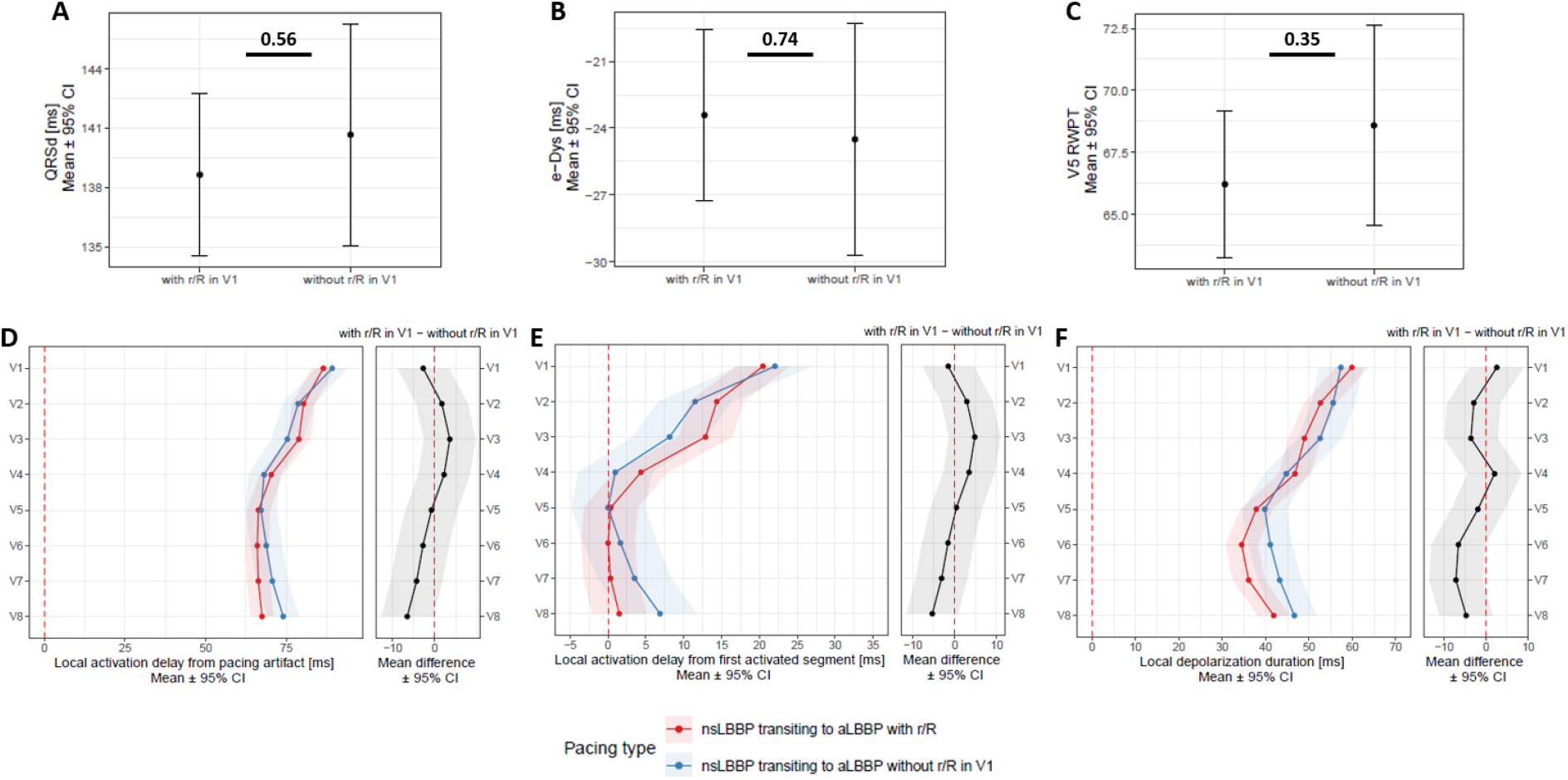
Panel A: QRSd, e-DYS (Panel B), V5RWPT (Panel C), delay of V1-V8 local activations from the pacing artifact (Panel D), local activations in V1-V8 (first activated segment was placed at 0 ms) (Panel E) and in Panel F local depolarization durations in V1–V8 between nsLBBP transitioning to aLBBP with late r/R and nsLBBP transitioning to aLBBP without the r/R morphology in the V1.

